# Evaluation of Cataract Surgery as a Cause of Astigmatism among Patients Undergoing Phacoemulsification and Extracapsular Cataract Extraction at KCMC Hospital

**DOI:** 10.1101/2024.04.04.24305351

**Authors:** Suzo Ambindwile Malakibungu, Andrew Makupa, William Makupa

## Abstract

**Background:** Despite the advance of cataract surgery astigmatism still occur after cataract surgery due to several reasons these include the preparation and closure of the surgical wound, the choice of suture material, and both intraoperative and postoperative manipulations in Phacoemulsification and Extracapsular cataract Etraction.

**Objectives:** To evaluate cataract surgery as cause of astigmatisms among patients undergoing phacoemulsification and extracapsular cataract extraction at eye department in KCMC hospital from Sept 2022 to April 2023

**Methods:** It is a clinic-based, prospective cohort study. Keratometric values and demographic data were collected for eligible patients who had undergone phacoemulsification and Extracapsular Cataract Extraction using a non-contact Auto Refkeratometer BARK-80.

**Results:** A total of 215 patients were recruited for the study. There were 129 (60%) females and 86 (40%) males. The mean age was 67.481 (SD 10.79years). A total 110 patients had undergone phacoemulsification and 105 had undergone Extracapsular Cataract Extraction (ECCE). 189 (87.91%), had a magnitude greater or equal (≥) to 0.5 post OP astigmatism. The mean corneal astigmatism among all patients undergone ECCE were 2.29D (SD 1.41 D) and all undergone PHACO were 0.95D (SD 0.79 D). The mean astigmatism among patients who had cataract surgery done by specialist were 1.19D (SD 0.09 D) and 2.41D (SD 0.16 D) were done by residents There was no astigmatism in 7 patients (3.26%), with-the-rule (WTR) in 68 patients (31.63%), against-the-rule (ATR) in 112 patients (52.09%) and oblique astigmatism (OA) in 28 patients (13.02%). The tendency of a gradual change from with the rule (WTR) to against the rule (ATR) astigmatism was noted as the age advanced.

**Conclusion:** The present study reveals the magnitude of astigmatism is higher in patients underwent Extra Capsular Cataract Extraction than Phacoemulsification in eye department at KCMC hospital and pre-existing astigmatism is a cause of surgical induced astigmatism if pre- operative astigmatism correction is not taken into consideration.

## Introduction

Astigmatism is one of the commonest refractive errors encountered during our clinical practice. Surgically induced astigmatism is the main obstacle to achieve good uncorrected visual acuity following cataract surgery(1) .Cataract is the leading cause of blindness in the world, and clinically significant astigmatism may affect up to approximately 20% of people undergoing cataract surgery(2).

A recent study reported the prevalence rate of astigmatism measuring 1 Dioptre (D) or more to be 23.9 % in adults(3). In India Over 40 % of the Indian patients undergoing cataract surgery have more than 1.0 D of corneal stigmatism and may benefit from the use of toric intraocular lenses(4).

More than 20 million of cataract surgery were expected to be done worldwide in 2015 (Review of ophthalmology, 2015). For routine cataract surgery, 41.3% of eyes had more than 1.00 D of corneal astigmatism and 11.6% had more than 2.00D. Females had more astigmatism than males (5).

Cornea astigmatism can be defined as a condition in which incident light rays are not refracted equally in all meridians. Thus, the refractive power of the eye varies with the orientation of the light rays. Usually, axes of greatest and least refractive powers can be determined. These are called the principal or major meridians. Astigmatism is said to be regular if the principal meridians are approximately 90” apart and irregular if they are not(6).

In the literature, the terms astigmatism “with-the- rule” and “against-the-rule” are frequently used. Regular astigmatism is said to be with-the-rule if the meridian with the greatest refractive power is near vertical in orientation or close to 90”and against-the-rule means that the greatest refractive power lies close to the horizontal, or near 180” (6).

The astigmatic change introduced due to the surgical treatment of cornea is called Surgically Induced Astigmatism (SIA) (7). Postoperative astigmatism results primarily from deformation of the cornea by surgery, it is related to the type, length, and location of the incision, and the suture closure technique (1). After a surgical procedure, the principal meridians may have any axial orientation, although certain operations, such as cataract surgery, often induce astigmatism with characteristic orientations (1).

At least 30000 consultations are done at KCMC eye department and an estimate of 1725 cataract surgeries is done each year. Clinically we see a lot of patients with post operative astigmatism and yet have not been studied the predators of astigmatism in these two procedures phacoemulsification and Extracapsular cataract extraction. The current study sought to bridge this gap, the study assessed the magnitude, predators and types of surgically induced astigmatism

## METHODOLOGY

### Study Design and Technique

This was a hospital based prospective cohort study, conducted at Kilimanjaro Christian Medical Center, Department of Ophthalmology from September 2022 to April 2023. Department of ophthalmology at Kilimanjaro Christian Medical Centre is a specialized eye care Centre in the northern Tanzania. It serves mainly Kilimanjaro, Arusha, Manyara and Tanga regions. It receives also referrals from the rest of the country and neighbouring countries. At least 30000 consultations are done at KCMC department of ophthalmology and an estimate of 1725 cataract surgeries is done each year.

### Study Site and Setting

The study was conducted at Kilimanjaro Christian Medical Center, Department of Ophthalmology. Eye department at Kilimanjaro Christian Medical Centre is a specialized eye care Centre in the northern Tanzania.

It serves mainly Kilimanjaro, Arusha, Manyara and Tanga regions. It receives also referrals from the rest of the country and neighbouring countries. At least 30000 consultations are done at KCMC eye department and an estimate of 1725 cataract surgeries is done each year.

Moreover, the eye department is a training institution for Ophthalmologists, Assistant Medical Officers Ophthalmology, Ophthalmic Nurses, Optometrists and Medical Students.

### Study population

Adult patients who underwent cataract surgery (Phacoemulsification and Extra Capsular Cataract Extraction) in eye department at KCMC hospital from 2022 to 2023.

### Eligibility criteria

#### Inclusion criteria

Patients who underwent phacoemulsification and Extra Capsular Cataract Extraction had age related cataract and were 40 years and above.

#### Exclusion criteria

Any pre-existing ocular conditions such as pterygium, corneal opacity, any patients with pre-existing corneal conditions, which could affect the Surgically Induced Astigmatism was excluded from our study

### Variables

Main outcomes were post-operative astigmatism.

Independent variables included Age, Pre-op keratometry readings, type of cataract surgery Surgeon (AMO, Resident, Ophthalmologist), type of incision, type of intraocular lens inserted recommended IOL power, inserted IOL power and suturing.

### Study Size

The minimum sample size was 140 patients calculated from the formula, where 70 patients for those Extra Capsular Cataract Extraction (ECCE) and other 70 patients for PHACOEMUSIFICATION cataract surgery. During study 215 subjects participated in the study, where 110 patients underwent ECCE and 105 underwent PHACOEMULSIFICATION.

### Data Collection tools and Procedure

All eligible patients were enrolled into the study after signs a consent form. All subjects underwent phacoemulsification and Extra Capsular Cataract Extraction (ECCE) had age related cataract and were 40 years and above. subjects were thoroughly examined at the clinic to rule out other conditions which affect cornea and cause astigmatism. Pre-operative keratometry was done by using a non-contact Auto Refkeratometer BARK-80, then subjects were taken to theatre for operation which were done by either specialists or trainees. Enrolled subjects were thoroughly examined on immediate postoperative day one and findings of uncollected visual acuity (UCVA), best collected visual acuity (BCVA), and keratometry was noted at the end of the 2^nd^ week and 6^th^ week follow up visits.

### Data Analysis

Database was created in SPSS version 25 then data was entered into the database. After data entry, cleaning and categorization of some variables done. Data were analysed by SPSS version 25. Descriptive analysis was done for demographic characteristics as well as for frequencies of independent variables. Independent sample t-test was used to compare means for continuous variables such as pre-operative astigmatism and post- operative astigmatism. In case of t-test, mean difference and 95% CI was calculated. Chi square was used for Predictors of Astigmatism in patients undergoing phacoemulsification and extracapsular cataract extraction

### Ethical clearance

This study was be approved by the Kilimanjaro Christian Medical University College Research and Ethics Review Committee (KCMU-CRERC) with clearance number PG 76/2022. Confidentiality of participants’ names, hospital personal details was be observed.

## Results

A total of 215 subjects underwent phacoemulsification with foldable intraocular lens (IOL) insertion and extracapsular cataract extraction with PMMA intraocular lens inserted. Findings showed that more phacoemulsification and extracapsular cataract extraction patients, 90 (41.86%), were aged 61-70 years and the mean age was 67.48 (SD: 10.79) years at baseline. More study participants 129 (60.00%) were females. The median cornea power was 44 diopters (IQR: 42.65-45.00). Pre-operative astigmatism were 95 subjects (44.19%) which was against the rule astigmatism. Hundred and ten subjects (51.16%) underwent Extracapsular Cataract Extraction (ECCE) surgery. Almost all (n=206, 95.81%) had no intra operation complication. Among those who had Extra Capsular Cataract Extraction, 68 (61.82%) had sutures. With reference to study participants, in the post-operated eye were 181 (84.19%) had normal BCVA and 189 (87.91%) had a magnitude greater or equal (≥) to 0.5 diopters post operative astigmatism. More results are shown in **Table 1**.

**Table 1.**
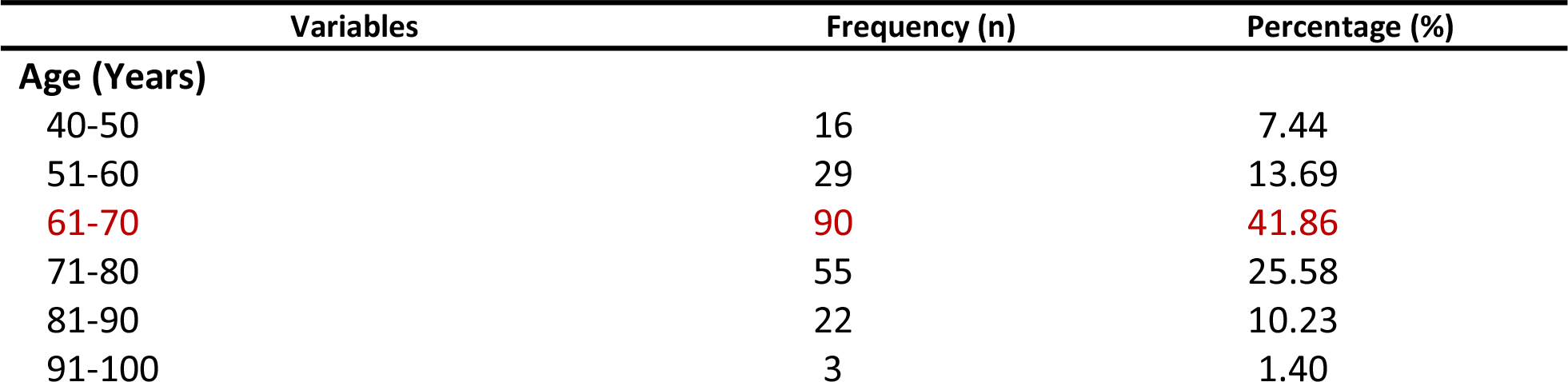
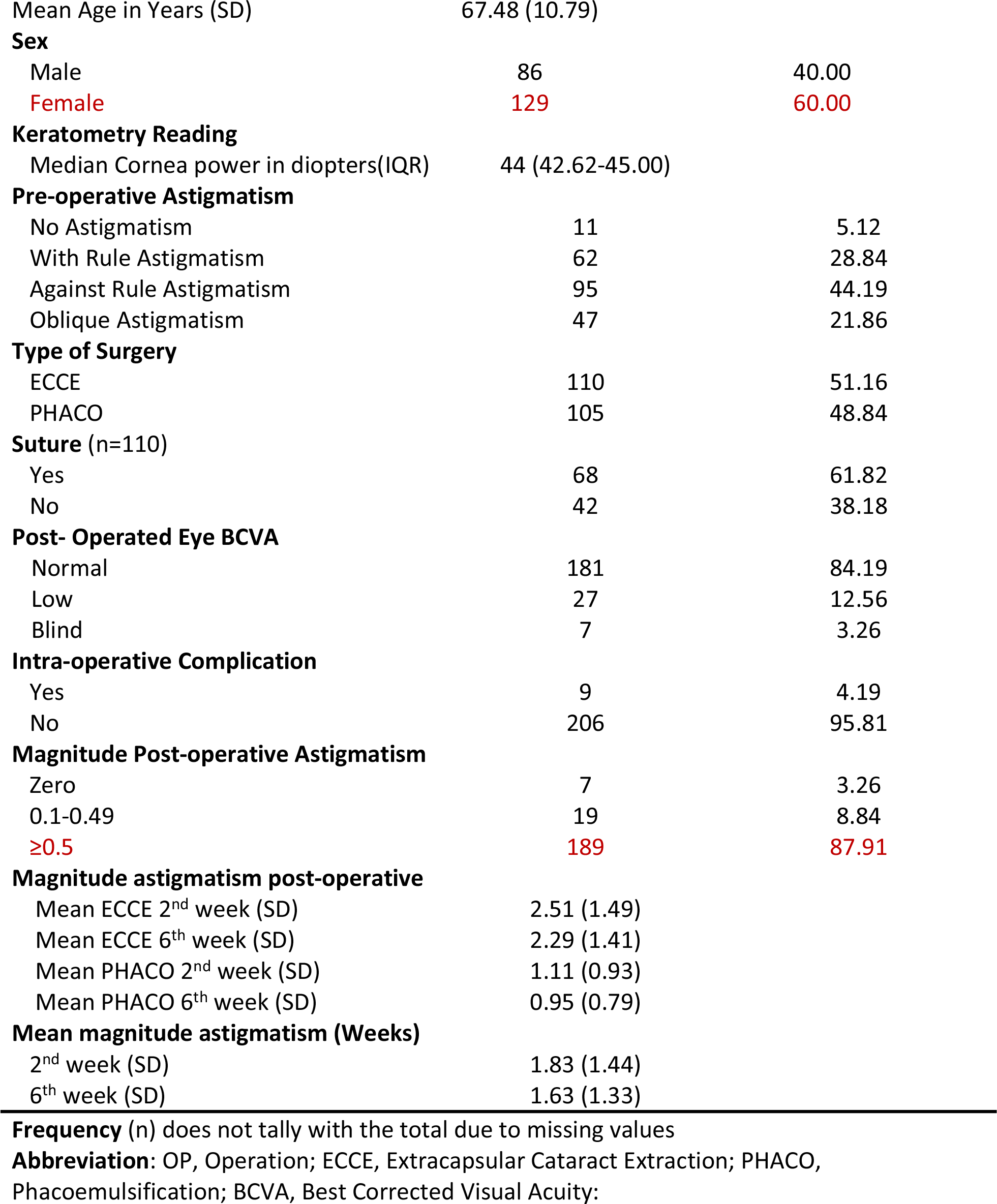
Socio-Demographic and Clinical characteristics in patients who underwent phacoemulsification and extracapsular cataract extraction (N=215)

| Variables | Frequency (n) | Percentage (%) |
| --- | --- | --- |
| **Age (Years)** |  |  |
| 40-50 | 16 | 7.44 |
| 51-60 | 29 | 13.69 |
| **61-70** | **90** | **41.86** |
| 71-80 | 55 | 25.58 |
| 81-90 | 22 | 10.23 |
| 91-100 | 3 | 1.40 |
| Mean Age in Years (SD) | 67.48 (10.79) |  |
| **Sex** |  |  |
| Male | 86 | 40.00 |
| Female | 129 | 60.00 |
| **Keratometry Reading** |  |  |
| Median Cornea power in diopters(IQR) | 44 (42.62-45.00) |  |
| **Pre-operative Astigmatism** |  |  |
| No Astigmatism | 11 | 5.12 |
| With Rule Astigmatism | 62 | 28.84 |
| Against Rule Astigmatism | 95 | 44.19 |
| Oblique Astigmatism | 47 | 21.86 |
| **Type of Surgery** |  |  |
| ECCE | 110 | 51.16 |
| PHACO | 105 | 48.84 |
| **Suture (n=110)** |  |  |
| Yes | 68 | 61.82 |
| No | 42 | 38.18 |
| **Post- Operated Eye BCVA** |  |  |
| Normal | 181 | 84.19 |
| Low | 27 | 12.56 |
| Blind | 7 | 3.26 |
| **Intra-operative Complication** |  |  |
| Yes | 9 | 4.19 |
| No | 206 | 95.81 |
| **Magnitude Post-operative Astigmatism** |  |  |
| Zero | 7 | 3.26 |
| 0.1-0.49 | 19 | 8.84 |
| ≥0.5 | 189 | 87.91 |
| **Magnitude astigmatism post-operative** |  |  |
| Mean ECCE 2<sup>nd</sup> week (SD) | 2.51 (1.49) |  |
| Mean ECCE 6<sup>th</sup> week (SD) | 2.29 (1.41) |  |
| Mean PHACO 2<sup>nd</sup> week (SD) | 1.11 (0.93) |  |
| Mean PHACO 6<sup>th</sup> week (SD) | 0.95 (0.79) |  |
| **Mean magnitude astigmatism (Weeks)** |  |  |
| 2<sup>nd</sup> week (SD) | 1.83 (1.44) |  |
| 6<sup>th</sup> week (SD) | 1.63 (1.33) |  |

**Frequency** (n) does not tally with the total due to missing values
**Abbreviation:** OP, Operation; ECCE, Extracapsular Cataract Extraction; PHACO, Phacoemulsification; BCVA, Best Corrected Visual Acuity:

**Table 2.** Proportion of Astigmatism by socio-demographic and clinical characteristics in patients who underwent phacoemulsification and extracapsular cataract extraction (N=215)

| Variable | Total | Astigmatism |  | Chi-square | P-value |
| --- | --- | --- | --- | --- | --- |
|  |  | Yes n (%)<br>208 (96.74) | No n (%)<br>7 (3.26) |  |  |
| **Suture** |  |  |  |  |  |
| Yes | 68 | 66 (97.06) | 2 (2.94) | 1.258 | 0.262 |
| No | 42 | 42 (100.00) | 0 (0.00) |  |  |
| **Type of surgery** |  |  |  |  |  |
| ECCE | 110 | 108 (98.2) | 2 (1.8) | 1.478 | 0.224 |
| PHACO | 105 | 100 (95.2) | 5 (4.8) |  |  |
| **Surgeon expertise** |  |  |  |  |  |
| Specialist | 137 | 130 (94.9) | 7 (5.1) | 4.119 | 0.042 |
| Resident | 78 | 78 (100.0) | 0 (0.0) |  |  |
| **Intraoperative complications** |  |  |  |  |  |
| Yes | 9 | 9 (100.0) | 199 (96.6) | 0.316 | 0.574 |
| No | 206 | 0 (0.0) | 7 (3.4) |  |  |
| **Pre-existing astigmatism** |  |  |  |  |  |
| No astigmatism | 11 | 9 (81.8) | 2 (18.2) | 8.504 | 0.037 |
| With rule | 62 | 61 (98.8) | 1 (1.6) |  |  |
| Against rule | 95 | 92 (96.8) | 3 (3.2) |  |  |
| Oblique | 47 | 46 (97.9) | 1 (2.1) |  |  |

**Abbreviation:** OP, Operation; ECCE, Extracapsular Cataract Extraction; PHACO, Phacoemulsification:

**Table 3.**
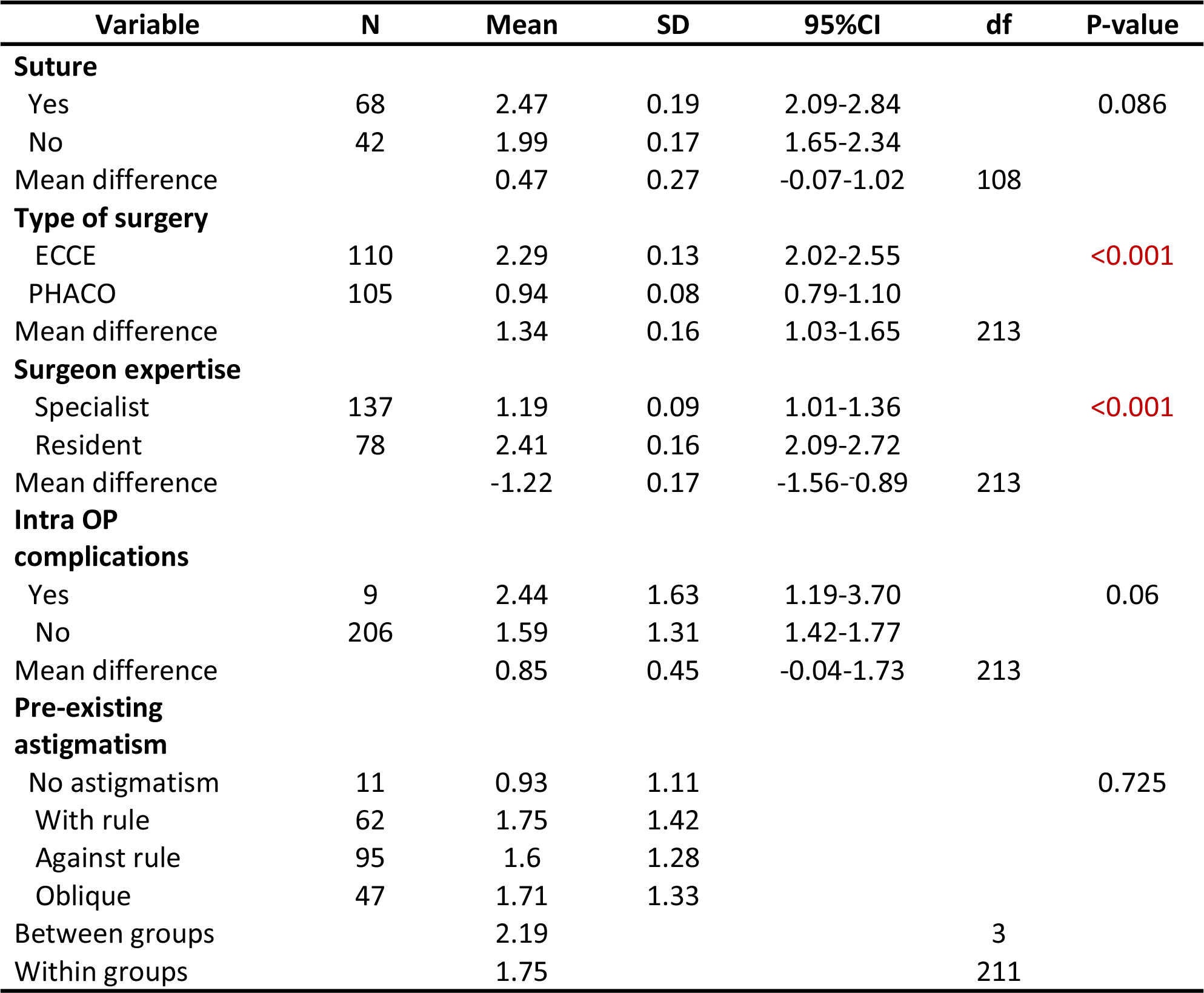
Predictors of astigmatism in patients underwent PHACO and ECCE (N=215)

### The proportion of Astigmatism among patients who underwent phacoemulsification and extracapsular cataract extraction

Among 215 subjects who underwent phacoemulsification and extracapsular cataract extraction with post-operative astigmatism, 16.74% had oblique astigmatism, 47.44% had against-rule astigmatism, 2.79% had no astigmatism, and 33.02% had with rule astigmatism within 2^nd^ week while 13.02% had oblique astigmatism, 52.09% had against-rule astigmatism, 3.26% had no astigmatism and 31.63% had with rule astigmatism within 6^th^ week as shown in **Figure 2**.

**Figure 1.**
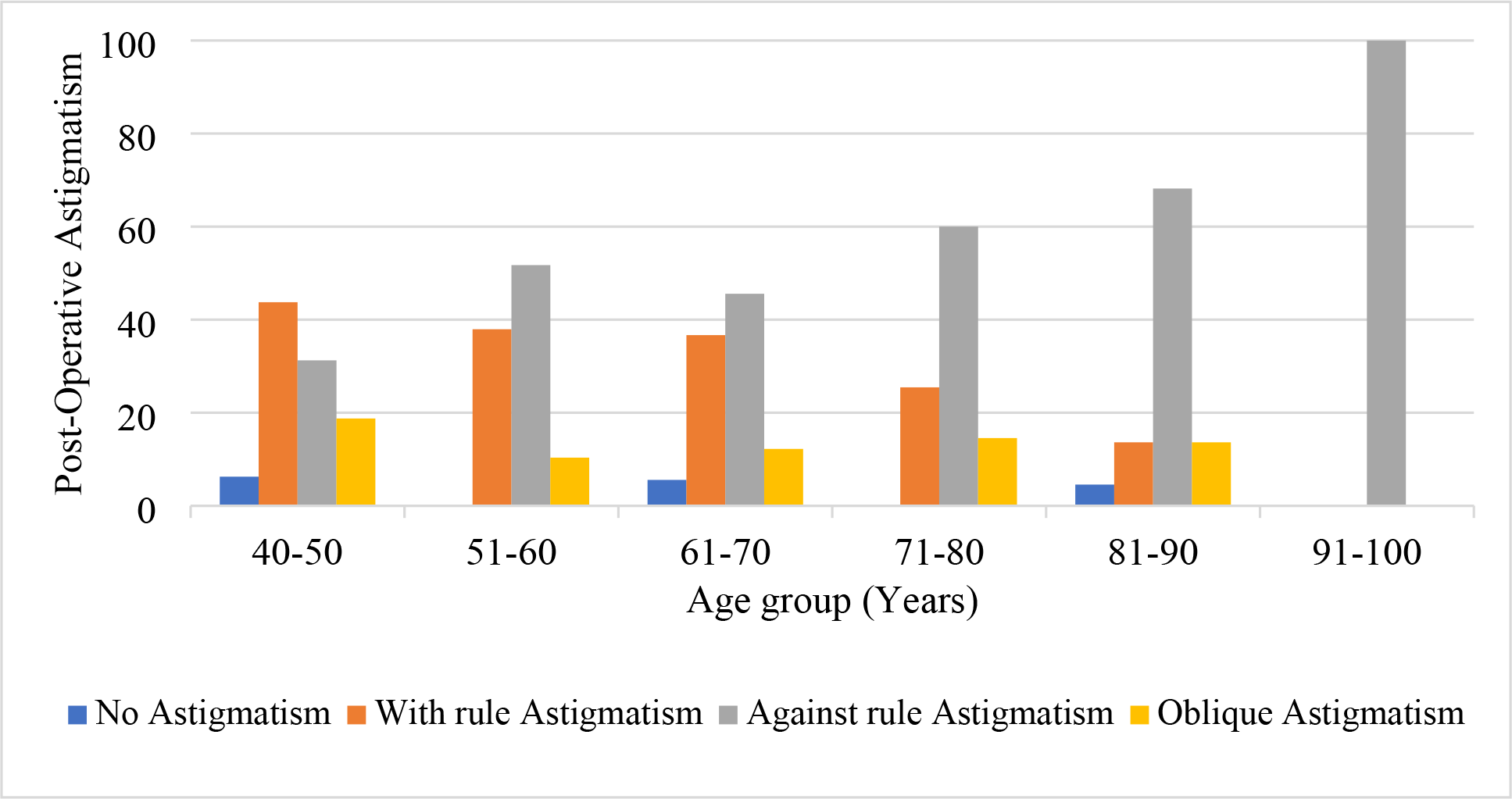
Proportion of Post-operative Astigmatism at 6th week by Age (N=215) p-value 0.332.

**Figure 2.**
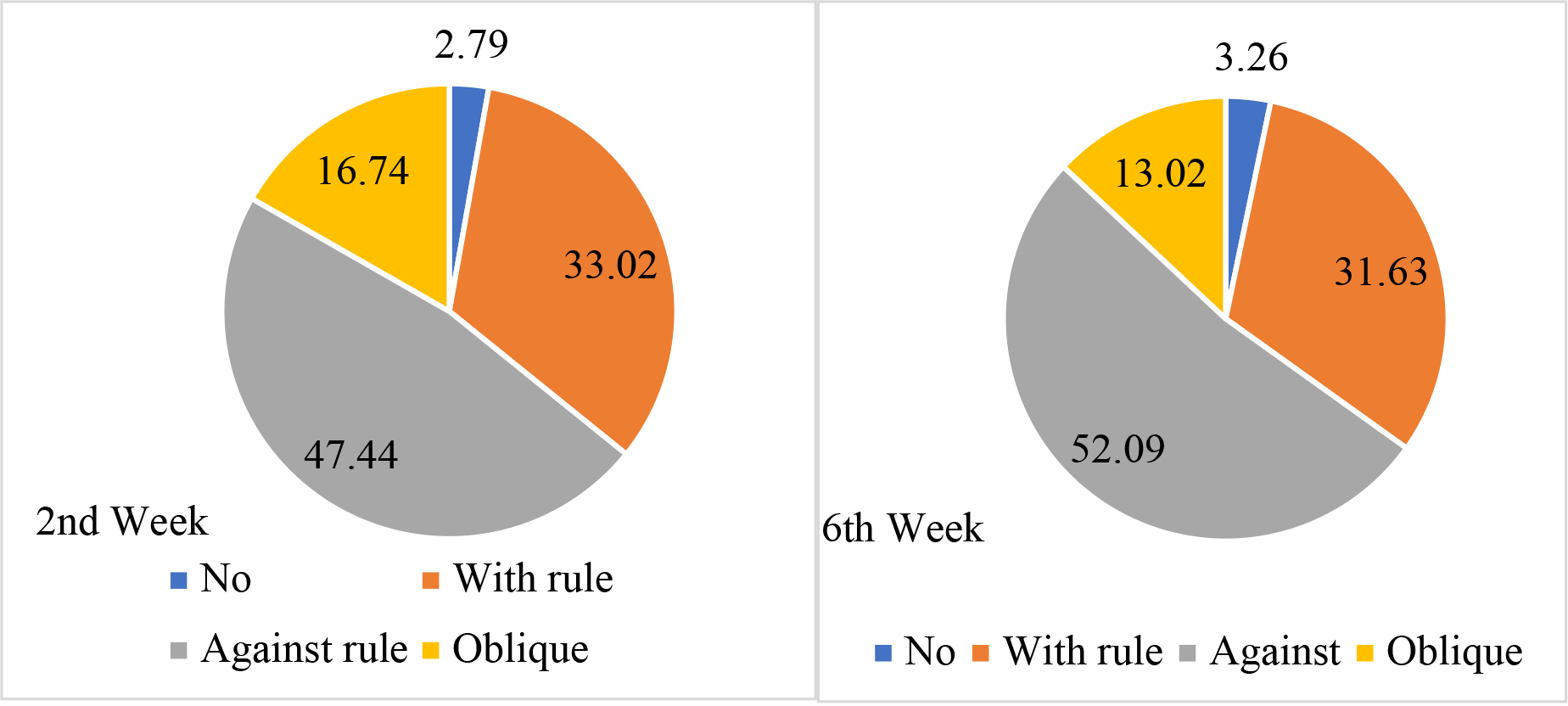
: Proportion of post-operative astigmatism among patients who went through phacoemulsification and extracapsular cataract extraction (N=215)

Among 215 subjects who underwent phacoemulsification and extracapsular cataract extraction with post-operative astigmatism, the mean magnitude of astigmatism in 2^nd^ week had significantly (p-value=0.038) increased with age and the mean magnitude of astigmatism in 6^th^ week had significantly (p-value=0.002) increased with age as shown in **Figure 3**.

**Figure 3:**
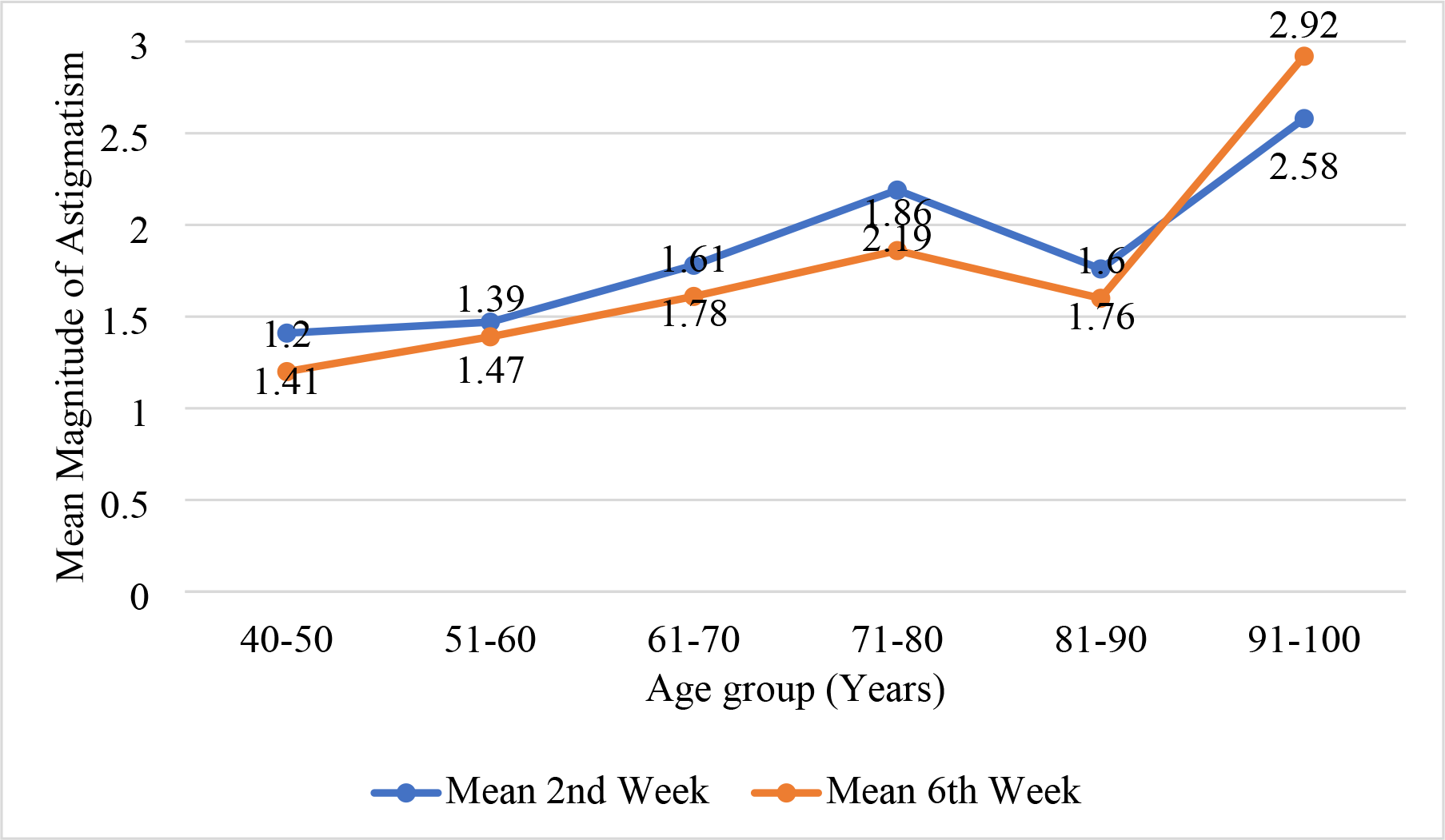
Mean Magnitude of Astigmatism (in dioptres) and age (in years)

## Discussion

The current study showed prevalence of visually significant cataract increases with age, patients aged above 60 years contributed to more than half the study population and 60% of patient were females. Similar results were reported in the previous study(8) and also to the study done by(9), mean age of 58.84±19. 57years.The sex distribution was 45% males and 55% females (9).

This study found that 87.91% of the patients had surgical induced astigmatism of more than 0.5 D and were thus the candidates for astigmatic correction. Similarly, to the study reported by(10), that the mean astigmatism postoperatively was more or equal to 0.5D in 90% of eyes. But some of previous study reported majority of patients had more than 1.00D. The difference could be attributed to eligibility criteria, age, racial factor, and methods of keratometry measurements (8).

The mean magnitude of astigmatism of this cohort study was not significantly different between any 2^nd^ week (1.83D SD 1.44) and 6^th^ week (1.63D SD 1.33). This suggests that on average the patient population was refractively stable at 2^nd^ week and remained so throughout the study duration. These data are consistent with previous reports showing early mean refractive stability in study populations (11).

Our current study has demonstrated that the induced corneal astigmatism following cataract surgery is considerably slighter after phacoemulsification than after extracapsular cataract extraction during at least the first six weeks postoperatively mean astigmatism in Extra Capsular Cataract Extraction and Phacoemulsification 2.29D (SD 1.41) and 0.95D (SD 0.79) respectively, our data was in accordance to (12), who reported the means and the standard deviation of Phacoemulsification post-operated astigmatism was 0.79 (SD 49) and Extracapsular Cataract Extraction post op astigmatism was 3.63 (SD 1.96) which indicated that the degree of astigmatism increases after extra capsular cataract extraction technique and also from (13), who reported a mean postoperative astigmatism was 0.91 ± 0.83 diopters in the Phacoemulsification group; 1.43 (SD 0.81) diopters in the Extra Capsular Cataract Extraction group.

In our study the mean magnitude of astigmatism was noted to increase with age and reduced in age group 40-60. A similar progressive increase in mean astigmatism with age has also been reported by (8), the mean magnitude of astigmatism was noted to decrease to a minimal in the age group of 40–60 years after which it was noted to increase progressively with age.

In the present study, against rule astigmatism was noted to be the most prevalent astigmatism contributing to 52.09%, with rule astigmatism 31.63%, oblique astigmatism 13.02% and no astigmatism 3.26% of the cases, which coincides with (8), found that after cataract surgery with the Rule astigmatism in 1062 eye (29.83%), Against Rule astigmatism in 1843 eyes (51.72%), oblique astigmatism in 555 eyes (15.59%) and no astigmatism in 99 eyes (2.7%).

The most common type of astigmatism in this study in the study population aged ≥61 years was against rule, which is in line with studies showing that astigmatism tends towards against rule as age (10). The current study reports the tendency of a gradual change from with the rule (WTR) to against the rule (ATR) astigmatism was noted as the age advanced, our data was in accordance to (8) who reported a gradual shift from WTR astigmatism to ATR astigmatism with an increase in age. This change was noted after 40 years of age.

Similar results have also been reported in previous studies on this subject (7). Various factors such as physiological changes in the corneal curvature as age advances, pressure from eyelids, pressure by intraocular pressure, and of the extraocular muscles have been anticipated to be responsible factors for changes in against rule and with rule with age.

Less astigmatism is not the only reason that patient’s vision is better without corrected visual acuity after careful phacoemulsification than after careful Extra Capsular Cataract Extraction. The larger incision and expression of the nucleus generally cause more corneal edema. Placement of the IOL in an intact capsule improves the accuracy of the postoperative refraction, and with rapid stabilization of the wound, the surgeon may prescribe spectacles soon after surgery, if any are needed.

This current study found that post -operated best collected visual acuity of operated eye 84.19% of patients achieved normal Best Collected Visual Acuity 6/12-6/18 while post op visual acuity of the better eye 92.09% of patients achieve normal Best Collected Visual Acuity at 6^th^ week postoperatively.

In this study reported mean magnitude of surgical induced astigmatism is 2.47D (SD 0.19) for patients had suture 9-0 nylon or 10-0 nylon after extracapsular cataract extraction. As most patients find it difficult to come for follow-up, this may be a better option as surgically induced astigmatism can be treated only if the patients present themselves for periodic refraction so that it can be picked up early and suture cutting carried out early (9). Though Drews 1995, cut no sutures and after 5years found no difference in astigmatism in wounds closed with10- 0nylonor11-0mersilene.Also Parkera and Clorfeine, 1989 found that the number of remaining10-0nylon sutures did not affect long term astigmatism (14).

Some investigators (14) have suggested that final postoperative astigmatism and refraction can be influenced by choice of suture material. Proper alignment and closure of the wound certainly are important to minimize postoperative complications, especially with large wounds Also, in this study found that sutureless manual cataract extraction the mean magnitude of surgical induced astigmatism is 1.99D (SD0.17), these results varied from (15), who reported Sutureless manual cataract extraction via subconjunctival limbus incision for mature cataract the mean surgical induced astigmatism was 0.62±0.41D. The difference could be attributed to different in sample size. However, carrying out sutureless extracapsular cataract extraction surgery, may reduce astigmatism almost completely as the amount of induced astigmatism is almost negligible when carried out carefully (9).

Findings in this study showed pre-existing is the predictor of post-operative astigmatism. Similarly, to previous study reported eyes with pre-existing astigmatism however have higher chances of being or remaining astigmatic after the surgery (9), it is varied from (14) study which shown that pre-operative astigmatism did not affect astigmatic change in their own series of Extra Capsular Cataract Extraction surgeries. However pre-existing astigmatism can be treated during surgery as shown by some studies (9), (16)

## Limitation

Our study has a few limitations such as not able to measure the size of the incision, not able to state type of incision, not able to measure site of incision from limbus and Lack of long term follow up.

## Conclusion

To conclude, the present study reveals the magnitude of astigmatism is higher in patients underwent Extra Capsular Cataract Extraction than Phacoemulsification in eye department at KCMC hospital and recorded more than 0.5 D of astigmatism in 87.91% of patients underwent cataract surgery. Corneal astigmatism was found to increase with age after 40 years and a shift from with rule astigmatism to Against rule astigmatism was noted with advancing age. With the improvement in the quality of healthcare and better age expectancy a greater number of patients would require quality vision following cataract surgery, which can only be achieved if pre-operative astigmatism correction is taken into consideration.

## Data Availability

data is available at KCMUCo for confidentiality

## Acknowledgment

I would like to highly appreciate the remarkable contributions and guidance of my Supervisors Dr. William Makupa and Dr. Andrew Makupa for adding value into this work. I also extend my sincere gratitude to everyone in the Department of ophthalmology for their valuable contributions. My appreciation extends to biostatistician Dr Winfrida Mwita, who donated her time to give her advices for work improvement. Last but not least, I convey special thanks to my family for their support and encouragement.

